# Using a polygenic score to account for genomic risk factors in a model to detect individuals with dilated ascending thoracic aortas

**DOI:** 10.1101/2023.09.06.23295145

**Authors:** John DePaolo, Gina Biagetti, Renae Judy, Grace J. Wang, John Kelly, Amit Iyengar, Nicholas J. Goel, Nimesh Desai, Wilson Y. Szeto, Joseph E. Bavaria, Michael G. Levin, Scott M. Damrauer

## Abstract

**Background:** Ascending thoracic aortic dilation is a complex trait that involves modifiable and non-modifiable risk factors and can lead to thoracic aortic aneurysm and dissection. Clinical risk factors have been shown to predict ascending thoracic aortic diameter. Polygenic scores (PGS) are increasingly used to assess clinical risk for multifactorial diseases. The degree to which a PGS can improve aortic diameter prediction is not known. In this study we tested the extent to which the addition of a PGS to clinical prediction algorithms improves the prediction of aortic diameter.

**Methods:** The patient cohort comprised 6,790 Penn Medicine Biobank (PMBB) participants with available echocardiography and clinical data linked to genome-wide genotype data. Linear regression models were used to integrate PGS weights derived from a large genome wide association study of thoracic aortic diameter in the UK biobank and were compared to the performance of the standard and a reweighted variation of the recently published AORTA Score.

**Results:** Cohort participants were 56% male, had a median age of 61 years (IQR 52-70) with a mean ascending aortic diameter of 3.4 cm (SD 0.5). Compared to the AORTA Score which explained 28.4% (95% CI 28.1% to 29.2%) of the variance in aortic diameter, AORTA Score + PGS explained 28.8%, (95% CI 28.1% to 29.6%), the reweighted AORTA score explained 30.4% (95% CI 29.6% to 31.2%), and the reweighted AORTA Score + PGS explained 31.0% (95% CI 30.2% to 31.8%). The addition of a PGS to either the AORTA Score or the reweighted AORTA Score improved model sensitivity for the identifying individuals with a thoracic aortic diameter ≥ 4 cm. The respective areas under the receiver operator characteristic curve for the AORTA Score + PGS (0.771, 95% CI 0.756 to 0.787) and reweighted AORTA Score + PGS (0.785, 95% CI 0.770 to 0.800) were greater than the standard AORTA Score (0.767, 95% CI 0.751 to 0.783) and reweighted AORTA Score (0.780 95% CI 0.765 to 0.795).

**Conclusions:** We demonstrated that inclusion of a PGS to the AORTA Score results in a small but clinically meaningful performance enhancement. Further investigation is necessary to determine if combining genetic and clinical risk prediction improves outcomes for thoracic aortic disease.

## Introduction

Thoracic aortic dilation can lead thoracic aortic aneurysm (TAA) development, a life-threatening condition. Unimpeded, TAA can progress to acute rupture or acute thoracic aortic dissection, which represent approximately 8% of all out-of-hospital cardiopulmonary arrests.^1^ For patients admitted to a hospital for emergent treatment of thoracic aortic aneurysm and dissection (TAAD), observed 30-day mortality is 39-49%.^2–5^ Despite the high mortality risk, most cases of ascending thoracic aortic dilation and subsequent aneurysm development are asymptomatic and identified incidentally.^6^ When recognized, asymptomatic ascending thoracic aortic aneurysms can be monitored with serial imaging and treated with aggressive blood pressure control and elective surgery, yielding significantly improved results when compared to emergency intervention.^7^

Across a range of cardiovascular diseases, polygenic scores (PGS) have become increasingly attractive as predictive tools to utilize in clinical settings to estimate individual level genetic risk for specific diagnoses.^8–10^ We and others have formulated PGSs to assess risk of increased ascending thoracic aortic diameter,^11,12^ and TAAD.^13^ GWAS have identified several genomic loci associated with both ascending thoracic aortic diameter and TAAD, and both traits are heritable, making PGS potentially useful for identifying individuals with enlarged aortas; however, the clinical utility of PGS remain poorly understood.

Recently, a clinical risk score to identify patients at elevated risk for ascending aortic dilation, referred to as the AORTA (aorta optimized regression for thoracic aneurysm) Score, was developed using cardiac magnetic resonance imaging (cMRI) measurements of ascending thoracic aortic diameter among individuals within the UK Biobank (UKB), and validated among subsets of individuals within the Framingham Heart Study (using computed tomography [CT] scans) and Mass General Brigham Biobank (using transthoracic echocardiography [TTE]).^14^ The present study validates the AORTA score within a separate diverse population, demonstrates the importance of creating individual population weighted values for specific factors in a predictive model, and shows the incremental benefit of including a PGS as a component of a combined clinical risk tool.

## Methods

### Study Population

Penn Medicine Biobank (PMBB) is composed of individuals recruited from across the Penn Medicine healthcare system. Each participant consents to linkage of electronic health records to biospecimens. A total of 44,297 volunteers have undergone genotyping per standard methodology as previously described.^15,16^ Among these individuals, 6,790 have at least one transthoracic echocardiography (TTE) study with a recorded measurement of ascending thoracic aortic diameter.

### Clinical Covariate Selection

We utilized the set of clinical factors identified in the original AORTA Score,^14^ including the presence of a diagnosis of hypertension, diabetes, and/or hyperlipidemia, sex, age, body mass index (BMI), weight, height, systolic and diastolic blood pressure, and pulse rate. We also included the previously reported series of interaction terms between the 11 clinical covariates.

### Prediction model creation

Using the clinical covariates and source code for the AORTA Score model (https://github.com/carbocation/genomisc), we derived predicted ascending thoracic aortic diameter values for PMBB participants and a linear regression model was fit using measured ascending thoracic aortic diameter as the dependent variable. The “AORTA Score + PGS” model was similarly constructed with PGS values included as a covariate for each individual in the PMBB. PGS scores were derived using weights from a previously published weighted allele PGS of ascending thoracic aortic diameter among UKB participants genetically similar to the European reference population.^12^ This PGS for ascending aortic diameter was constructed using 89 autosomal independently significant SNPs from individuals genetically similar to the European reference population in the UKB. Principal components of ancestry were derived for all PMBB participants and included as covariates in the AORTA Score + PGS model.

Three additional comparator models were created to assess the performance of the both the AORTA Score and the AORTA Score + PGS among PMBB individuals. First, to improve model performance by leveraging local population health characteristics, a reweighted version of the AORTA Score with clinical weights derived from individuals within PMBB, subsequently referred to as the “reweighted AORTA Score,” was created without consideration of PGS or principal components of ancestry. Second, this model was expanded to include the PGS plus the first five PCs of ancestry, which is subsequently referred to as the “reweighted AORTA Score + PGS.” Third, an age plus sex model that included both PGS and the first five principal components of ancestry, subsequently referred to as the “Age + Sex + PGS model,” was also analyzed to determine the effect of genetic factors without considering clinical risk factors.

### Model evaluation

To limit over-fitting, performance measures are reported using 5 repeats of 10-fold cross-validation. This repeated cross-validation splits the data evenly 10 times, holds out one of 10 folds for model assessment while the data across the remaining nine folds are used to fit the model. This entire process is repeated five times, resulting in 50 separate model fits/tests. Final resampling estimates of performance average each cross-validated replicate with 95% credible intervals.

Model performance and calibration were evaluated by several methods including *R*-squared (RSQ) and root-mean-square error (RMSE) using *yardstick* (v1.2.0),^17^ linear regression effect estimate (coefficient of the slope of the linear model), and intercept (calibration). Predicted and residual aortic diameters were plotted against actual measurements. Bland-Altman plots were created for each individual model.^18^

Between-model comparisons were systematically made using Bayesian analysis of variance (ANOVA) function in the *tidyposterior* package in R (v1.0.0) that relied upon random intercept modeling to fully account for resampling by assuming that individual resamples only effect the model by changing the intercept.^19^ We generated posterior distributions of RSQ and RMSE values with corresponding credible intervals to compare individual model performance. This allowed the derivation of probability that the proportion of the comparative model RSQ and RMSE posteriors are greater than zero; that is, the probability that the estimated performance of the two models is different. Posterior distributions were also used to compute the probability of being practically equivalent or significant with a practical effect size of +/-2%, also known as the Region of Practical Equivalence (ROPE) estimate.^20^

### Score Thresholding and Confusion Matrix Analysis

A range of score threshold values between three and four were tested in confusion matrices to assess the appropriate linear model cutoff to optimize sensitivity and specificity and predict ascending thoracic aortic diameter ≥ 4 cm. For each respective linear model, score threshold values were set at intervals increasing by five percent, representing the fifth percent highest score to the 50^th^ percent highest score. These thresholds were used to identify the true-positives, false-positives, true-negatives, and false-negatives at any given cutoff value compared to the actual values, and score sensitivity and specificity were analyzed.

### Binary model construction and Decision Curve Analysis

Logistic regression models were used to predict the ascending thoracic aortic diameter ≥ 4 cm based on AORTA Score covariates and PGS. To test prediction performance, the area under the receiver operator characteristic curve (AUROC) was calculated for each model using the *pROC* package in R (v1.18.1).^21^ Ninety-five percent confidence intervals for the AUROC were calculated using the *ci.auc* function.

Clinical net benefit was assessed for each logistic regression model using decision curve analysis, an analytic instrument to test the benefit of a diagnostic tool in the presence of competing harms and benefits.^22,23^ Decision curve analysis was performed on each model with “net benefit” defined as cases of ascending thoracic aortic diameter ≥ 4 cm identified for screening imaging per 100 individuals. These analyses generate a plot of net benefit as a function of “threshold probability” at which a clinician or individual considers the potential benefit versus harm of undergoing TTE imaging. For instance, if a clinician were to say that there is no harm to unnecessary screening, the “threshold probability” would equal 0%. If, however, a clinician believed screening represented an unnecessary risk no matter what could be found on subsequent imaging, the “threshold probability” would equal 100%. TTE imaging has minimal risks, as it is non-invasive, in-expensive, and has no radiation exposure; therefore, the threshold probability assessed in this study was 0-25%. Decision curve analysis was performed using *dcurves* (v0.4.0).^24^

### Statistical Analyses

Statistical analysis was performed in R version 4.2.1 (R Foundation for Statistical Computing, Vienna, Austria). The *Tidymodels* R package (v1.0.0) was used for all model cross-validation analysis.^25^ Throughout this investigation, Bayesian statistics were used to derive posterior probabilities and credible intervals of individual model performance statistics.

## Results

### Clinical Characteristics

A total of 6790 PMBB participants were included. Forty-four percent of the individuals were female, and the median age was 61 years (IQR 52 – 70) [**Table 1**]. The mean ascending aortic diameter was 3.4 cm, and there were 830 individuals (12%) with an ascending aortic diameter ≥ 4 cm. Seventy-four percent of individuals had a history of hypertension, 63% had a history of hyperlipidemia, and 33% had a history of diabetes. Compared to the UKB training cohort from which the AORTA Score was derived, the PMBB cohort had increased prevalence of co-morbidities and larger average ascending thoracic aortic diameter (**Table 1**).

**Table 1.** Clinical characteristics of Penn Medicine Biobank individuals compared to the UKB training cohort. UKB training dataset data taken from Pirruccello, et al.^14^.

| UK Biobank and Penn Medicine Biobank Participant Characteristics |  |  |  |
| --- | --- | --- | --- |
|  |  | UKB Training Dataset | PMBB Dataset |
|  |  | (N = 30,018) | (N = 6790) |
| Sex, N (%) | Female | 15635 (52.1) | 2990 (44.0) |
|  | Male | 14383 (47.9) | 3800 (56.0) |
| Ancestry, N (%) | European |  | 4448 (65.5) |
|  | African |  | 2116 (31.2) |
|  | Other |  | 226 (3.3) |
| Age in years, median (IQR) |  | 65.1 (58.6-70.6) | 61.0 (52.3-69.7) |
| Medical history, N (%) | Hypertension | 9009 (30.0) | 5013 (73.8) |
|  | Hyperlipidemia | 6380 (21.3) | 4252 (62.6) |
|  | Diabetes | 876 (2.9) | 2204 (32.5) |
| Weight in kg, median (IQR) |  | 74.0 (64.7-84.2) | 87.0 (73.6-100.5) |
| Height in cm, median (IQR) |  | 169 (162-176) | 172 (164.0-180.0) |
| BMI, median (IQR) |  | 25.7 (23.4-28.5) | 29.3 (25.2-33.5) |
| HR in beats/ minute, median (IQR) |  | 67.6 (61.0-75.0) | 77.0 (69.8-84.2) |
| Blood pressure in mmHg, median (IQR) | Systolic | 138 (127-150) | 127.8 (119.9-135.8) |
|  | Diastolic | 78.5 (72.2-85.0) | 73.9 (69.2-78.7) |
| Ascending Aortic Diameter in cm | Mean (SD) | 3.18 (0.35) | 3.37 (0.50) |
|  | Median (IQR) | 3.15 (2.93-3.40) | 3.3 (2.95-3.65) |
| Aortic diameter 4+ cm, N (%) |  | 696 (2.3) | 830 (12.2) |

### PMBB Model Calibration and Performance

The performance of the standard AORTA Score and each model variation were utilized to estimate the aortic diameter for each PMBB participant within the cohort. For a 1 cm estimate for ascending thoracic aortic diameter using the AORTA Score, the measured aortic diameter was 1.08 cm (95% confidence interval [CI] 1.07 cm to 1.15 cm, *P*<0.001) suggesting a significant underestimation of aortic diameter; this was not seen with any of the other models (**Table S1**). Predicted values for individual models were plotted against measured and residual values (**Figure S1**). In Bland-Altman plots, the 95% limits of agreement of the AORTA Score 0.63 cm to -1.04 cm, suggesting the model underestimated large diameters more than it overestimated small diameters, and generally conservative errors (**Figure S2**). Bland-Altman plots for each of the other comparator models demonstrated more balanced 95% limits of agreement with consistently conservative errors.

Individual model RSQ and RMSE demonstrated that the reweighted AORTA Score + PGS was the best performing model, and that the addition of PGS a conferred a small performance improvement on both the AORTA Score and the reweighted AORTA Score (**Table 2**). Compared to the AORTA Score which explained 28.4% (95% CI 28.1% to 29.2%) of the variance in aortic diameter, AORTA Score + PGS explained 28.8%, (95% CI 28.1% to 29.6%), the reweighted AORTA score explained 30.4% (95% CI 29.6% to 31.2%), and the reweighted AORTA Score + PGS explained 31.0% (95% CI 30.2% to 31.8%).

**Table 2:** R-squared (RSQ) and Root-Mean-Square Error (RMSE) with 95% CI for each model variation based on 5 repeats of 10-fold cross-validation analysis.

| Model RSQ and RMSE |  |  |
| --- | --- | --- |
| Model | RSQ (95% CI) | RMSE (95% CI) |
| AORTA Score | 0.284 (0.276 to 0.292) | 0.426 (0.423 to 0.430) |
| AORTA Score + PGS | 0.288 (0.281 to 0.296) | 0.425 (0.421 to 0.429) |
| Reweghted AORTA Score | 0.304 (0.296 to 0.312) | 0.420 (0.417 to 0.424) |
| Reweghted AORTA Score + PGS | 0.310 (0.302 to 0.318) | 0.419 (0.415 to 0.422) |
| Age + Sex + PGS | 0.225 (0.216 to 0.234) | 0.444 ( 0.440 to o.448) |

CI = Credible Interval; PGS = polygenic score.

To determine specific inter-model performance differences, we applied Bayesian principles to resampled data from PMBB to generate a set of posterior probabilities and credible intervals (CI) for the RSQ and RMSE of each model for ANOVA analysis between models.^25^ The differences in RSQ between each of AORTA Score versus AORTA Sore + PGS (RSQ mean difference 0.002, 95% CI -0.002 to 0.005) and reweighted AORTA Score versus reweighted AORTA Score + PGS (RSQ means difference 0.002, 95% CI -0.001 to 0.006) suggested that the addition of the PGS to either model incrementally improve explanation of variance **(Table S2, Figure 1**). Comparable results were observed when investigating direct comparison of RMSE between these models suggesting PGS also improves model accuracy (**Table S3, Figure S3**). However, neither marginal improvement was practically different using a region of practical equivalence (ROPE) of 2% (**Tables S4 and S5**). These results suggest that the addition of the PGS modestly improves model explanation of variance (RSQ) and accuracy (RMSE) independent of reweighting in a statistically significant but not clinically meaningful manner.

**Figure 1.**
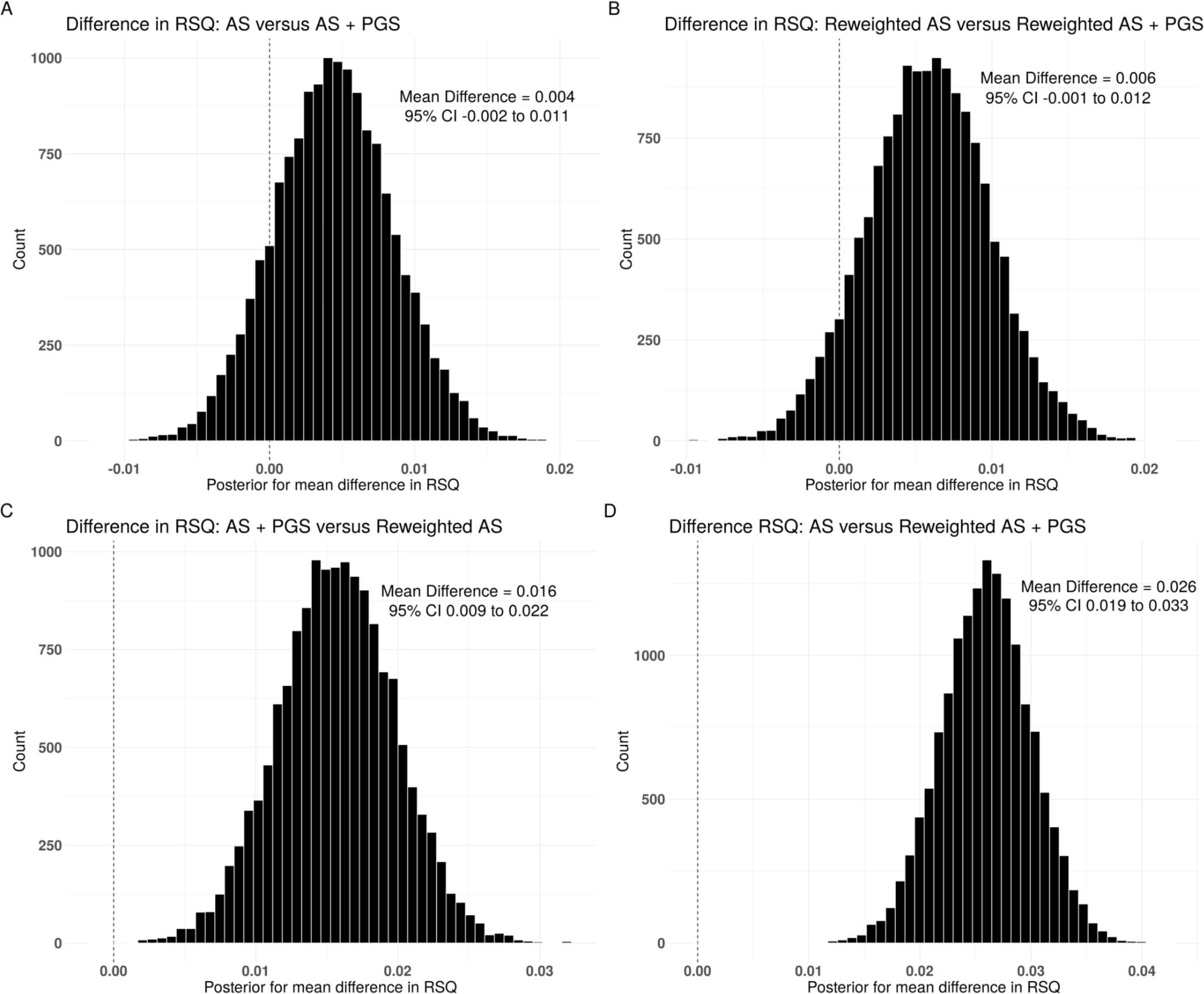
Between-model comparison of difference in explanation of variance (RSQ). Cross-validated posterior distribution for the difference in RSQ between (A) standard AORTA Score and AORTA Score + PGS; (B) reweighted AORTA Score and reweighted AORTA Score + PGS; (C) AORTA Score + PGS and reweighted AORTA Score; (D) standard AORTA Score and reweighted AORTA Score + PGS. Between-model mean difference and 95% credible intervals (CIs) in top right corner of each plot. PGS = polygenic score; AS = AORTA Score; RSQ = *R*-squared.

As the AORTA Score variables are commonly assessed clinically and require no additional testing, if performance improvements can be made by simple reweighting, a PGS may not be necessary as it may require additional cost/testing. To assess if the addition of a PGS to the standard AORTA Score conferred a similar performance improvement compared as model reweighting, we directly compared AORTA Score + PGS and the reweighted AORTA Score. The reweighted AORTA Score had superior explanation of variance (RSQ mean difference 0.016, 95% CI 0.009 to 0.022) and accuracy (RMSE mean difference 0.005, 95% CI 0.002 to 0.008) [**Tables S2 and S3, Figures 1 and S3**]. However, neither difference achieved likely practical significance (**Table S4 and S5**). These findings suggest that model reweighting based on local baseline health characteristics improves AORTA Score performance marginally more than the addition of a PGS.

### Assessing Model Clinical Utility

To assess the clinical performance of each model in predicting individuals with aortic diameters ≥ 4 cm, a clinically meaningful threshold at which serial imaging would be recommended, we performed logistic regression analysis using the covariates from the AORTA score, and PGS where indicated, and analyzed receiver operator characteristic curves (ROC) and area under the ROC (AUROC) (**Figure 2**). The AORTA Score + PGS (AUROC = 0.771, 95% CI 0.756 to 0.787), reweighted AORTA Score (AUROC = 0.780 95% CI 0.765 to 0.795), and reweighted AORTA Score + PGS (AUROC = 0.785, 95% CI 0.770 to 0.800) each better predicted ascending thoracic aortic diameter ≥ 4 cm compared to the standard AORTA Score (AUROC = 0.767, 95% CI 0.751 to 0.783) and Age + Sex + PGS (AUROC = 0.737, 95% CI 0.720 to 0.753). Taken together, these results suggest that the addition of the PGS to the standard and reweighted AORTA Scores improves the ability of either model to predict aortic diameter greater than 4 cm, and that the optimal model is the reweighted AORTA Score + PGS.

**Figure 2.**
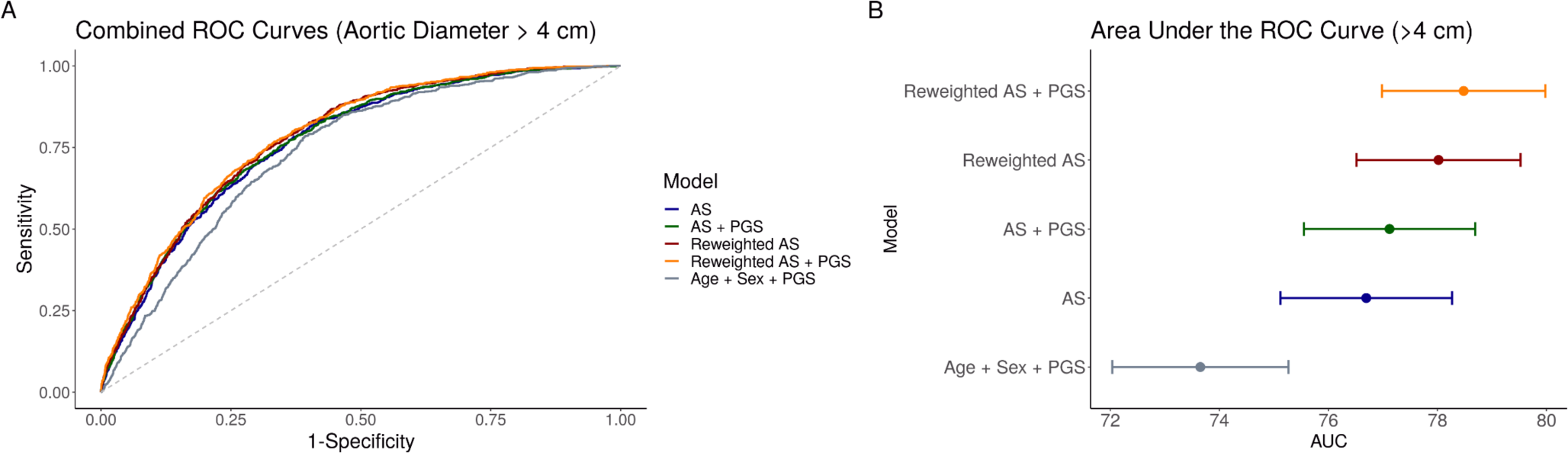
Logistic model receiver operator characteristic curve (ROC) and area under the ROC curve to predict ascending thoracic aortic diameter ≥ 4 cm. A) Receiver operator characteristic curve (ROC) for each of the five models analyzed in the PMBB cohort. B) Area under the ROC curves with error bars demonstrating 95% confidence intervals. PGS = polygenic score; AS = AORTA Score.

To investigate whether PGS integration provides net clinical benefit to the AORTA Score, we performed decision curve analyses (DCA) using each logistic regression model. DCA allows the assessment of net clinical benefit which equates to the number of true positives screened less the number of false positives screened at a specific threshold probability.^22^ In this evaluation, the screening test is a transthoracic echocardiogram, a non-invasive and relatively inexpensive procedure. Using a risk threshold of 0-25%, each of the five models evaluated performs better than the “Screen All” or “Screen None” strategies (**Table S6, Figure 3**). At the threshold probability of 25% for example, the net benefit of the reweighted AORTA Score + PGS logistic regression model was 1.5 cases per 100 individuals screened.

**Figure 3:**
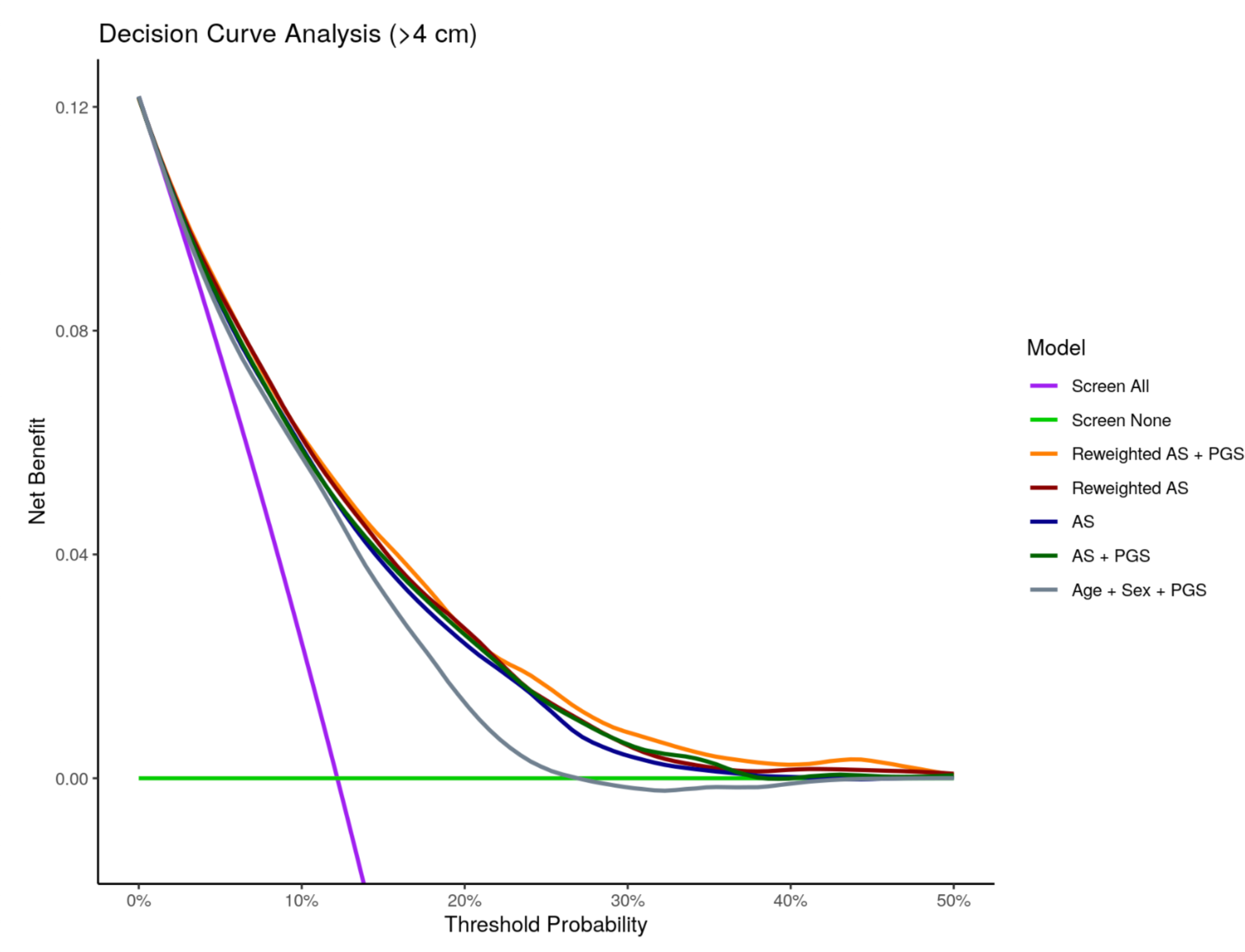
Individual model decision curve analysis. Decision curve analysis demonstrated clinical net benefit for each model plotted against threshold probability with units representing cases identified per individual screened at a specific threshold probability using a range of 0-50%. Results are compared to a model that would screen all individuals (“Screen All”) or screen no one (“Screen None”). PGS = polygenic score; AS = AORTA Score.

This net benefit was increased compared to the reweighted AORTA Score logistic regression (net benefit of 1.4 cases per 100 individuals screened), AORTA Score + PGS logistic regression (1.4 cases per 100 individuals screened), the standard AORTA Score logistic regression (1.0 cases per 100 individuals screened), and the Age + Sex + PGS logistic regression (0.2 cases per 100 individuals screened). These data suggest that the addition of the PGS and reweighting the AORTA Score based on local clinical risk factors may slightly enhance the overall clinical utility of the baseline AORTA Score model (approximately 1 additional case per 200 individuals screened), and provides further evidence that the addition of a PGS incrementally improves model performance.

As a sensitivity analysis we constructed confusion matrices using each linear regression model at different score thresholds and compared model sensitivity and specificity to compare optimal model thresholds that could result in maximized clinical utility to discern individuals who would benefit from screening (**Tables S7 and S8**). Thresholds were set every five percent from the fifth percentile score to the 50^th^ percentile score. These scores were then utilized as cutoff values for predicting a case (ascending thoracic aortic diameter ≥ 4 cm) versus controls. Across thresholds, the reweighted AORTA Score + PGS was consistently the best performing model; for instance, using the 10^th^ percentile score in every model, the reweighted AORTA Score + PGS had a sensitivity of 29.8% compared to 27.7% for the reweighted AORTA Score, 27.0% for the AORTA Score + PGS, 26.5% for the AORTA Score, and 22.4% for the Age + Sex + PGS. The addition of the PGS frequently contributed to improved model sensitivity supporting our observation that the addition of a PGS incrementally improves model performance.

## Discussion

Using a published predictive model of enlarged ascending aortic diameter, we demonstrated that the addition of a PGS based on the largest available GWAS of ascending thoracic aortic diameter modestly improved standard AORTA Score and reweighted AORTA Score performance by several measures including the explanation of variance (RSQ) and accuracy (RMSE), however these improvements in performance fell within a range of practical equivalence of 2%. The addition of PGS also marginally improved the predictive capacity to identify aortic diameter ≥ 4 cm as determined by AUROC, sensitivity, and overall net benefit in decision curve analyses. Notably, in the sensitivity assessment the addition of the PGS incrementally improved model performance at a range of thresholds suggesting that the benefit is not limited to a narrow cohort with elevated risk. We also demonstrated that using PMBB participant clinical data to reweight the AORTA Score improved model performance more than the addition of the PGS alone, and that the combination of reweighting and PGS created the best performing model.

Currently, there are no meaningful recommendations for screening individuals for dilated ascending thoracic aortas who lack a history of familial disease. Our findings are instructive when considering how to clinically implement a screening model for ascending thoracic aortic aneurysm. Both the addition of genetic information and reweighting the covariates improved AORTA Score model fit and corrected aortic diameter underestimation observed with the standard AORTA Score model in PMBB. Though the predictive improvement in AORTA Score from the addition of the PGS and/or from reweighting was marginal, both confusion matrix and decision curve analysis suggest that these changes will consistently benefit clinical decision-making more than the current strategy (i.e., screen none) by identifying individuals at risk for aortic dilation who would benefit from a non-invasive TTE. Furthermore, the addition of the PGS, covariate reweighting, and the combination of both consistently perform better than the standard AORTA Score within the threshold probability ranges assessed.

AORTA Score covariate reweighting based on local health characteristics easily aggregable from electronic health record (EHR) platforms may offer a more expedient strategy for rapid, optimal clinical implementation of AORTA Score as it avoids reliance on genetic data that are not ubiquitously available. However, in clinical settings where large biobanks have been established, integration of the PGS and reweighting the AORTA Score would most successfully identify individuals who warrant screening TTEs. These centers of care may even consider implementing a program similar to Recall by Genotype (RbG) initiatives where all enrolled individuals are screened using the AORTA Score + PGS and those at elevated risk are contacted and offered TTEs. As genetic data becomes increasingly available due to the rapid speed and availability of whole genome sequencing, integration of a model accounting for both local health characteristics and genetic data may become more universally applicable with the potential to more accurately identify individuals who warrant screening.

Notably, the modest improvements currently attributable to the PGS are derived from the largest GWAS available for ascending thoracic aortic diameter. This GWAS included 38,694 individuals, 2-3% of whom had aortic diameters ≥ 4 cm. Compared to other more powerful PGS available in other cardiovascular diseases, this is a relatively small cohort of individuals from which to derive PGS weights. While ascending thoracic aortic diameter was found to be highly heritable in this GWAS (63%, 95% CI 60–67),^12^ even our best-performing model explained only a modest proportion of variance in aortic diameter, and substantially less variability than the anticipated heritability. As GWAS sizes expand and understanding of the genetic underpinnings of ascending aortic diameter and aneurysmal degeneration improve, the AORTA Score enhancement attributable to PGS inclusion will likely continue to grow.

### Limitations

This study has several limitations. First, both the AORTA Score and the PGS for ascending thoracic aortic dilation were derived from the UK Biobank whose participants are largely healthier and less diverse than most of the major urban centers within the US from which regional biobanks like the PMBB are constructed. For instance, not only do PMBB participants have a larger ascending thoracic aortic diameter to begin with (mean 3.36 cm compared to 3.18 cm), but the PMBB training cohort had a higher median BMI (29.3 compared to 25.7), rates of hypertension (73.9% compared to 30.0%), hyperlipidemia (62.9% compared to 21.3%), and diabetes (32.5% compared to 2.9%). While these factors were considered in the reweighted AORTA Score, it remains unclear how these health factors would impact broader implementation of the AORTA Score, and how the underlying genetic risks for these conditions may impact aortic dilation.

Additionally, this study was limited to a single biobank. The effect of the PGS may be limited as a function of sample size and power. Sensitivity and specificity calculations also depend highly on disease prevalence, which is relatively elevated compared to UKB. Further investigation among other cohorts will contribute to assessing the benefit of including the PGS as a covariate within the AORTA Score.

### Conclusions

Our findings suggest that the reweighted AORTA score model using PMBB data and incorporating PGS weights is the best performing linear regression model to predict enlarged ascending thoracic aortic diameter. Independent of reweighting, the addition of genomic covariates modestly improved overall performance of the standard AORTA Score and the reweighted AORTA Score suggesting that utilization of genetic data may enhance clinical outcomes when determining individuals who warrant screening for ascending thoracic aortic dilation. Further research is needed to determine if similar cohorts have improved model performance with the addition of a PGS as a covariate, and whether that improvement translates to better clinical outcomes among a diverse population with differing levels of baseline health.

## Data Availability

All data produced in the present study are available upon reasonable request to the authors.

## Acknowledgements

We thank the participants of the Penn Medicine Biobank. The PMBB is supported by Perelman School of Medicine at University of Pennsylvania, a gift from the Smilow family, and the National Center for Advancing Translational Sciences of the National Institutes of Health under CTSA award number UL1TR001878. J.D. is supported by the American Heart Association (23POST1011251). M.G.L. is supported by the Institute for Translational Medicine and Therapeutics of the Perelman School of Medicine at the University of Pennsylvania, the NIH/NHLBI National Research Service Award postdoctoral fellowship (T32HL007843), the Measey Foundation, and the Doris Duke Foundation. S.M.D. is supported by funding from the Department of Veterans Affairs Office of Research (IK2-CX001780).

