## Supplementary material for "Using a polygenic score to account for genomic risk factors in a model to detect individuals with dilated ascending thoracic aortas": Tables S1-S8, Figures S1-S3

**Affiliations:**
[1] Department of Surgery, Perelman School of Medicine, University of Pennsylvania, Philadelphia, PA 19104, USA.
[2] Division of Vascular Surgery, Department of Surgery, Perelman School of Medicine, University of Pennsylvania, Philadelphia, PA 19104, USA.
[3] Division of Cardiovascular Surgery, Department of Surgery, Perelman School of Medicine, University of Pennsylvania, Philadelphia, PA 19104, USA.
[4] Cardiovascular Institute, Department of Medicine, Perelman School of Medicine, University of Pennsylvania, PA 19104, USA.
[5] Corporal Michael Crescenz VA Medical Center, Philadelphia, PA 19104, USA.
[6] Department of Genetics, Perelman School of Medicine, University of Pennsylvania, Philadelphia, PA 19104, USA.
*co-last authors

**Table S1.** Model calibration of the linear regression effect estimate (coefficient of the slope of the linear model), intercept (calibration) with 95% CI for each model.

| Model Calibration and Testing Analysis | | | | | | |
|---|---|---|---|---|---|---|
| Model | Effect Estimate (cm) | 95% CI | P-value | Intercept (cm) | 95% CI | P-value |
| AORTA Score | 1.08 | 1.07 to 1.15 | <0.001 | -0.14 | -0.27 to 0.00 | 0.044 |
| AORTA Score + PGS | 1.00 | 0.96 to 1.04 | <0.001 | 0.00 | -0.13 to 0.13 | 1.000 |
| Reweighted AORTA Score | 1.00 | 0.96 to 1.04 | <0.001 | 0.00 | -0.12 to 0.12 | 1.000 |
| Reweighted AORTA Score + PGS | 1.00 | 0.97 to 1.04 | <0.001 | 0.00 | -0.12 to 0.12 | 1.000 |
| Age + Sex + PGS | 1.00 | 0.96 to 1.04 | <0.001 | 0.00 | -0.15 to 0.15 | 1.000 |

CI = Credible Interval; PGS = polygenic score.

**Table S2.** Probability of difference and mean difference between cross-validated posterior distributions of RSQ between select model variations.

| RSQ Posterior Probability Differences between Models | | | | | |
|---|---|---|---|---|---|
| Reference Model | Comparator Model | Probability Different | Mean Difference | Lower CI | Upper CI |
| AORTA Score | AORTA Score + PGS | 0.858 | 0.004 | -0.002 | 0.011 |
| Reweighted AORTA Score | Reweighted AORTA Score + PGS | 0.925 | 0.006 | -0.001 | 0.012 |
| AORTA Score + PGS | Reweighted AORTA Score | 1.000 | 0.016 | 0.009 | 0.022 |
| AORTA Score | Reweighted AORTA Score + PGS | 1.000 | 0.026 | 0.019 | 0.033 |

CI = Credible Interval; PGS = polygenic score; RSQ = *R*-squared.

**Table S3**: Probability of difference and mean difference between cross-validated posterior distributions of RMSE between select model variations.

**RMSE Posterior Probability Differences between Models**

| Reference Model | Comparator Model | Probability Different | Mean Difference | Lower CI | Upper CI |
|---|---|---|---|---|---|
| AORTA Score | AORTA Score + PGS | 0.756 | 0.001 | -0.002 | 0.004 |
| Reweighted AORTA Score | Reweighted AORTA Score + PGS | 0.827 | 0.002 | -0.001 | 0.005 |
| AORTA Score + PGS | Reweighted AORTA Score | 0.994 | 0.005 | 0.002 | 0.008 |
| AORTA Score | Reweighted AORTA Score + PGS | 1.000 | 0.008 | 0.005 | 0.011 |

CI = Credible Interval; PGS = polygenic score; Score; RMSE = Root-mean-square error.

**Table S4.** Probability of practical differences between individual model posterior distribution for the mean difference in RSQ using region of practical equivalence (ROPE) +/- 2%.

| **RSQ Practical Differences between Models** | | | |
|---|---|---|---|
| Reference Model | Comparator Model | Practically Equivalent | Practically Positive Difference |
| AORTA Score | AORTA Score + PGS | 1.000 | 0.000 |
| Reweighted AORTA Score | Reweighted AORTA Score + PGS | 1.000 | 0.000 |
| AORTA Score + PGS | Reweighted AORTA Score | 0.855 | 0.145 |
| AORTA Score | Reweighted AORTA Score + PGS | 0.073 | 0.927 |

PGS = polygenic score; RSQ = *R*-squared.

**Table S5:** Probability of practical differences between individual model posterior distribution for the mean difference in RMSE using region of practical equivalence (ROPE) +/- 2%.

| RMSE Practical Differences between Models | | | |
|---|---|---|---|
| Reference Model | Comparator Model | Practically Equivalent | Practically Positive Difference |
| AORTA Score | AORTA Score + PGS | 1 | 0 |
| Reweighted AORTA Score | Reweighted AORTA Score + PGS | 1 | 0 |
| AORTA Score + PGS | Reweighted AORTA Score | 1 | 0 |
| AORTA Score | Reweighted AORTA Score + PGS | 1 | 0 |

PCs = first 5 principal components; PGS = polygenic score; AS = AORTA Score.

**Table S6:** Decision curve analysis demonstrating net benefit for each model within a threshold probability of 0-25% compared to hypothetical models where all individuals are screened ("Screen All") or no individuals are screened ("Screen None").

| Decision Curve Analysis (Threshold Probability 0-25%) | | | | | | |
|---|---|---|---|---|---|---|
| Model | 0% Threshold Probability | 5% Threshold Probability | 10% Threshold Probability | 15% Threshold Probability | 20% Threshold Probability | 25% Threshold Probability |
| AS | 0.122 | 0.081 | 0.057 | 0.037 | 0.022 | 0.010 |
| AS + PGS | 0.122 | 0.081 | 0.057 | 0.038 | 0.023 | 0.014 |
| Reweighted AS | 0.122 | 0.083 | 0.060 | 0.039 | 0.022 | 0.014 |
| Reweighted AS + PGS | 0.122 | 0.084 | 0.059 | 0.038 | 0.025 | 0.015 |
| Age + Sex + PRS | 0.122 | 0.081 | 0.057 | 0.032 | 0.012 | 0.002 |
| "Screen All" | 0.122 | 0.076 | 0.024 | -0.033 | -0.098 | -0.171 |
| "Screen None" | 0.000 | 0.000 | 0.000 | 0.000 | 0.000 | 0.000 |

Results are presented in net benefit units meaning cases identified per individual screened at a specific threshold probability. PGS = polygenic score.

**Table S7**. Model sensitivity calculated from confusion matrices using threshold percentage scores among individuals within PMBB with higher (fifth percentile) or lower (50th percentile) thresholds at which a case (ascending thoracic aortic diameter ≥ 4 cm) is predicted.

| Model Sensitivity at a Range of Score Thresholds | | | | | |
|---|---|---|---|---|---|
| Threshold (%) | AS | AS + PGS | Reweighted AS | Reweighted AS + PGS | Age + Sex + PGS |
| 5 | 14.7 | 15.8 | 15.7 | 16.8 | 11.3 |
| 10 | 26.5 | 27.0 | 27.7 | 29.8 | 22.4 |
| 15 | 39.5 | 39.3 | 39.4 | 41.9 | 32.0 |
| 20 | 48.6 | 49.8 | 50.5 | 50.1 | 40.7 |
| 25 | 56.6 | 55.0 | 58.1 | 59.7 | 49.8 |
| 30 | 63.7 | 63.5 | 65.0 | 66.6 | 59.4 |
| 35 | 70.1 | 68.2 | 71.2 | 71.9 | 66.0 |
| 40 | 75.1 | 75.7 | 77.0 | 77.8 | 71.7 |
| 45 | 80.9 | 80.1 | 82.0 | 82.1 | 79.1 |
| 50 | 84.3 | 84.7 | 86.9 | 86.5 | 83.4 |

**Table S8**. Model specificity calculated from confusion matrices using threshold percentage scores among individuals within PMBB with higher (fifth percentile) or lower ($50^{th}$ percentile) thresholds at which a case (ascending thoracic aortic diameter ≥ 4 cm) is predicted.

| Model Specificity at a Range of Score Thresholds | | | | | |
|---|---|---|---|---|---|
| Threshold (%) | AS | AS + PGS | Reweighted AS | Reweighted AS + PGS | Age + Sex + PGS |
| 5 | 96.3 | 96.9 | 96.5 | 96.6 | 95.9 |
| 10 | 92.3 | 92.8 | 92.4 | 92.7 | 91.7 |
| 15 | 88.4 | 88.8 | 88.4 | 88.7 | 87.4 |
| 20 | 84.0 | 84.3 | 84.2 | 84.2 | 82.9 |
| 25 | 79.4 | 79.9 | 79.6 | 79.8 | 78.5 |
| 30 | 74.7 | 74.7 | 74.9 | 75.1 | 74.1 |
| 35 | 69.9 | 69.6 | 70.0 | 70.1 | 69.3 |
| 40 | 64.9 | 64.9 | 65.1 | 65.2 | 64.5 |
| 45 | 60.0 | 59.9 | 60.1 | 60.2 | 59.8 |
| 50 | 54.8 | 54.8 | 55.1 | 55.1 | 54.6 |

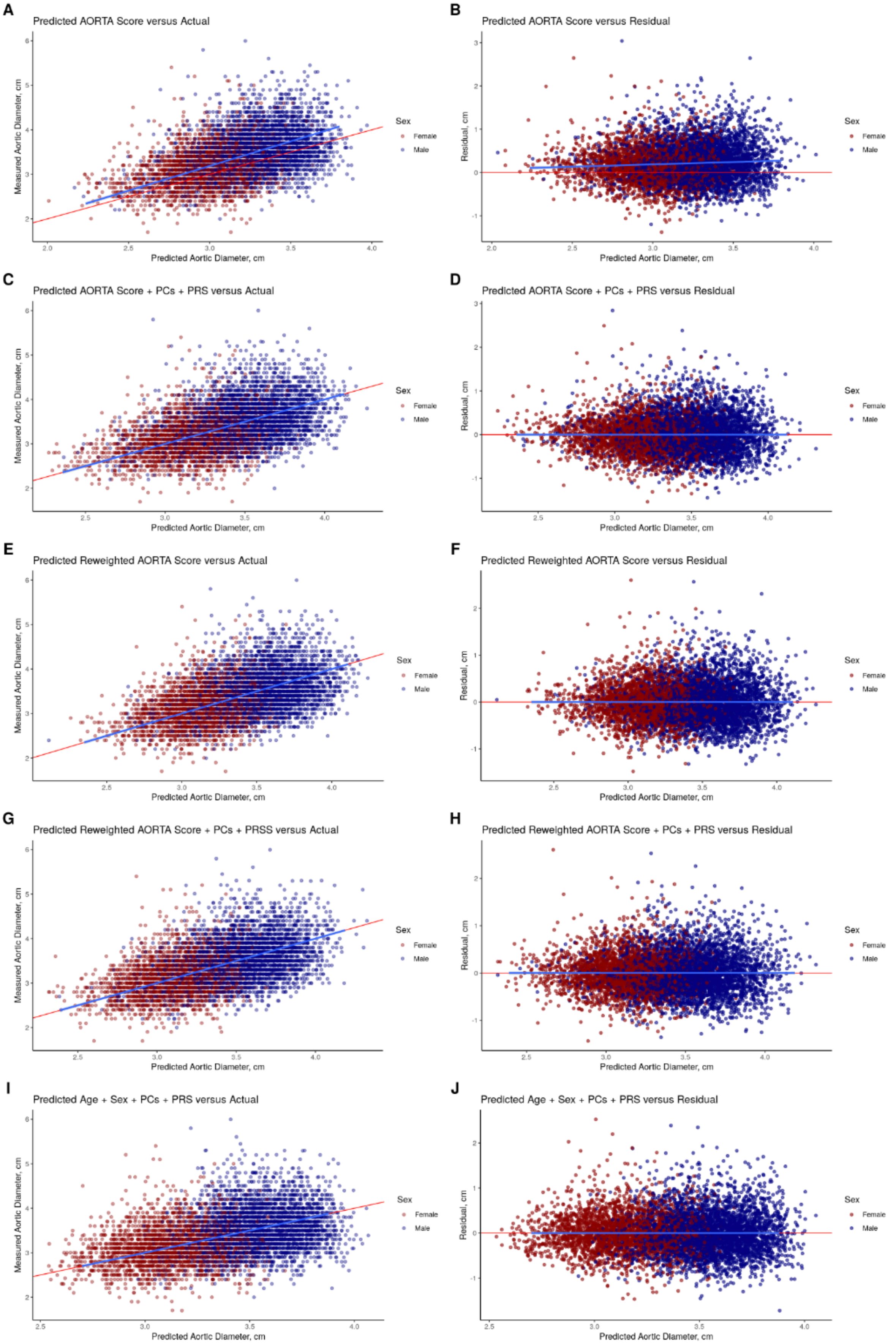

A
Predicted AORTA Score versus Actual
B
Predicted AORTA Score versus Residual
C
Predicted AORTA Score + PCs + PRS versus Actual
D
Predicted AORTA Score + PCs + PRS versus Residual
E
Predicted Reweighted AORTA Score versus Actual
F
Predicted Reweighted AORTA Score versus Residual
G
Predicted Reweighted AORTA Score + PCs + PRSS versus Actual
H
Predicted Reweighted AORTA Score + PCs + PRS versus Residual
I
Predicted Age + Sex + PCs + PRS versus Actual
J
Predicted Age + Sex + PCs + PRS versus Residual
Measured Aortic Diameter, cm
Residual, cm
Predicted Aortic Diameter, cm
Sex
Female
Male

**Figure S1.** Predicted versus actual plots for each individual model (A, C, E, G, and I) stratified by sex. Residual versus actual plots for each individual model (B, D, F, H, and J) stratified by sex. For each panel, a point represents one of 6790 PMBB participants, the blue line indicates the slope of the average of each model with the grey region representing 95% CIs for the average, and the red line indicates the ideal curve for each model.

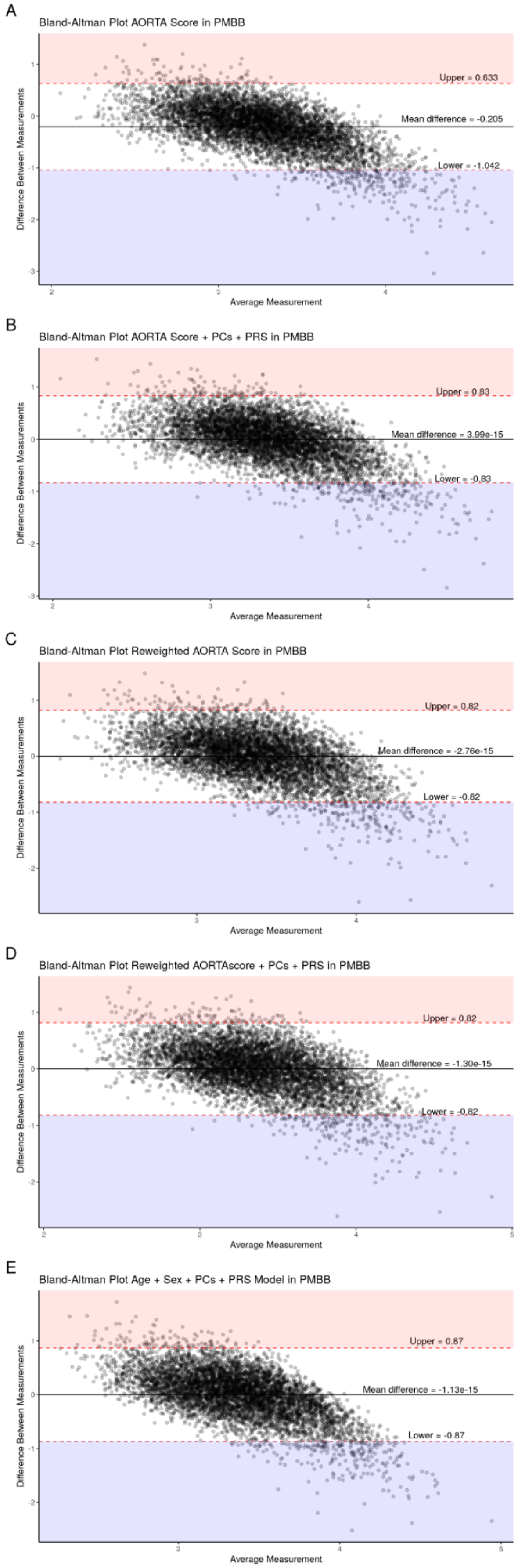
A
Bland-Altman Plot AORTA Score in PMBB
Upper = 0.633
Mean difference = -0.205
Lower = -1.042
Difference Between Measurements
Average Measurement
B
Bland-Altman Plot AORTA Score + PCs + PRS in PMBB
Upper = 0.83
Mean difference = 3.99e-15
Lower = -0.83
Difference Between Measurements
Average Measurement
C
Bland-Altman Plot Reweighted AORTA Score in PMBB
Upper = 0.82
Mean difference = -2.76e-15
Lower = -0.82
Difference Between Measurements
Average Measurement
D
Bland-Altman Plot Reweighted AORTAscore + PCs + PRS in PMBB
Upper = 0.82
Mean difference = -1.30e-15
Lower = -0.82
Difference Between Measurements
Average Measurement
E
Bland-Altman Plot Age + Sex + PCs + PRS Model in PMBB
Upper = 0.87
Mean difference = -1.13e-15
Lower = -0.87
Difference Between Measurements
Average Measurement

**Figure S2.** Bland-Altman plots for the agreement between each individual model predicted ascending thoracic aortic diameter and measured aortic diameter among the 6790 individuals in the PMBB cohort. For each point, the x-axis represents the mean of the model score and the measured aortic diameter, and the y-axis and represents the different between the predicted and measured diameter. The mean difference as well as he lower and upper bounds of the 95% limits of agreement with regions of over (red) and under (blue) estimate of each individual model score.

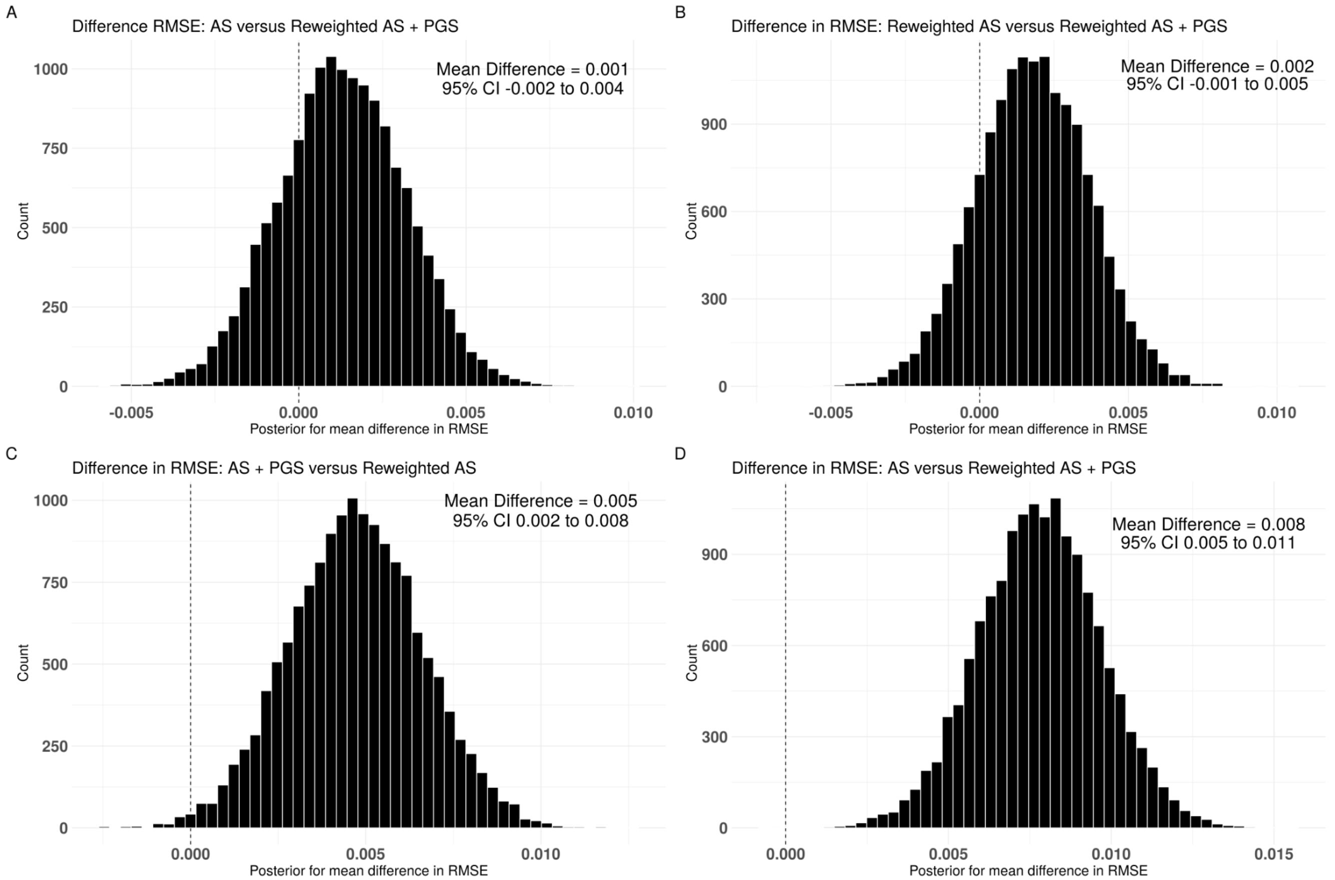


**Figure S3**. **Between-model comparison of difference in root-mean-square error (RMSE).** Cross-validated posterior distribution for the difference in RMSE between (A) standard AORTA Score and AORTA Score + PGS; (B) reweighted AORTA Score and reweighted AORTA Score + PGS; (C) AORTA Score + PGS and reweighted AORTA Score; (D) standard AORTA Score and reweighted AORTA Score + PGS. Between-model mean difference and 95% credible intervals (CIs) in top right corner of each plot. PGS = polygenic score; AS = AORTA Score; RMSE = root-mean-square error.
